# Geographic and temporal trends in fentanyl-detected deaths in Connecticut, 2009-2019

**DOI:** 10.1101/2022.07.26.22278052

**Authors:** Haidong Lu, Forrest W. Crawford, Gregg S. Gonsalves, Lauretta E. Grau

**Affiliations:** Department of Epidemiology of Microbial Diseases, Yale School of Public Health, New Haven, CT, USA; Public Health Modeling Unit, Yale School of Public Health, New Haven, CT, USA; Department of Biostatistics, Yale School of Public Health, New Haven, CT, USA; Department of Statistics & Data Science, Yale University, New Haven, CT, USA; Department of Ecology & Evolutionary Biology, Yale University, New Haven, CT, USA; Yale School of Management, New Haven, CT, USA; Yale Law School, New Haven, CT, USA

**Keywords:** fentanyl, overdose, opioid crisis, Bayesian disease mapping, spatiotemporal analysis

## Abstract

**Purpose:** Since 2012 fentanyl-detected fatal overdoses have risen from 4% of all fatal overdoses in Connecticut to 82% in 2019. We aimed to investigate the geographic and temporal trends in fentanyl-detected deaths in Connecticut during 2009-2019.

**Methods:** Data on the dates and locations of accidental/undetermined opioid-detected fatalities were obtained from Connecticut Office of the Chief Medical Examiner. Using a Bayesian space-time binomial model, we estimated spatiotemporal trends in the proportion of fentanyl-detected deaths.

**Results:** During 2009-2019, a total of 6,632 opioid deaths were identified. Among these, 3,234 (49%) were fentanyl-detected. The modeled spatial patterns suggested that opioid deaths in northeastern Connecticut had higher probability of being fentanyl-detected, while New Haven and its neighboring towns and the southwestern region of Connecticut, primarily Greenwich, had a lower risk. Model estimates also suggested fentanyl-detected deaths gradually overtook the preceding non-fentanyl opioid-detected deaths across Connecticut. The estimated temporal trend showed the probability of fentanyl involvement increased substantially since 2014.

**Conclusion:** Our findings suggest that geographic variation exists in the probability of fentanyl-detected deaths, and areas at heightened risk are identified. Further studies are warranted to explore potential factors contributing to the geographic heterogeneity and continuing dispersion of fentanyl-detected deaths in Connecticut.

## INTRODUCTION

The three waves of the American opioid crisis (i.e., prescription opioids, heroin, and synthetic opioids other than methadone) continue to be a significant public health emergency in the United States (US).^1^ Beginning in late 2013, synthetic opioid overdoses (chiefly illicitly produced fentanyl and fentanyl analogs) have dominated the ongoing third wave of the opioid crisis.^2^ Illicit drug supply is the key driver of the current opioid crisis.^3^ Illicitly manufactured fentanyl and fentanyl analogs, which are less costly to produce and distribute (but are 50 to 100 times more potent than morphine),^4^ have increased in the US drug market.^5^ Reports from the US Drug Enforcement Administration highlighted the dramatic increase in drug seizures that tested positive for fentanyl, from 4,642 in 2014 to 98,954 in 2019 in the US.^6,7^ The national rate of drug overdose deaths involving fentanyl increased dramatically from 0.5 per 100,000 persons in 2011 to 5.9 per 100,000 in 2016.^8^ From 2014 to 2015, overdose deaths attributed to synthetic opioids (primarily illicitly manufactured fentanyl) increased by 72% to nearly 10,000.^9^ Most recently, the rate of drug overdose deaths involving synthetic opioids rose by 27%, from 9.0 per 100,000 in 2017 to 11.4 in 2019.^10,11^ In 2018, fentanyl and its analogs were detected in approximately two thirds of opioid deaths.^12^

Abrupt changes in illicit drug supply may have contributed to the heightened risk of opioid overdoses since 2013 and the influx of fentanyl.^3^ Examining the spatiotemporal trends of fentanyl-detected deaths may help public health researchers and policymakers better understand geographic and temporal variation in fentanyl availability. While several studies have been conducted to investigate the small-area geographic distribution of fentanyl-detected overdose deaths^13,14^, to our knowledge, no research has explored whether the fentanyl-detected overdose deaths simply take over the rising overall opioid overdoses in the same geographic region or whether fentanyl has established new regions of risk for overdose where, previously, overdoses were less common.

To further inform overdose prevention and intervention efforts, we investigated the geographic and temporal trends of fentanyl-detected overdose deaths at the city/town level in Connecticut, a state that is highly affected by the opioid crisis. We examined whether spatiotemporal distributions of fentanyl-detected overdose deaths covary with overall trends of opioid overdose deaths, and whether the rising tide of fentanyl-detected overdose deaths follows a geographic pattern different from that of the preceding wave of opioid overdose deaths. Our town-level estimates will help support the decision-making of the local health agencies (organized by town or health districts consisting of several towns) in Connecticut on public health and surveillance efforts to address the opioid crisis.

## METHODS

### Study Sample and Data

We obtained data on all opioid-detected overdose deaths from the records of the Connecticut Office of the Chief Medical Examiner (OCME), including the cause and manner of death, toxicological test results in available specimens, demographic information (i.e., age, sex, race/ethnicity), and the addresses of residence, injury, and/or death for each case.^15,16^ In the OCME records, “opioid detection” in an overdose death means that either an opioid was listed in the cause of death or that a quantifiable amount of an opioid was present in the toxicological test report.

All opioid-detected overdose deaths from 2009 to 2019 in Connecticut that the OCME determined to be of accidental or undetermined manner were included in this analysis. Non-residents in Connecticut were excluded. We dichotomized the opioids detected in the overdose deaths (i.e., fentanyl-detected versus non-fentanyl-detected). Other substances of interest included heroin/morphine, pharmaceutical opioids (i.e., di-hydrocodeine, hydromorphone, hydrocodone, hydromorphone, oxymorphone, and tramadol), methadone/buprenorphine, benzodiazepines, cocaine, ethanol, methamphetamine, xylazine, mitragynine (kratom), and gabapentin. This research project has been reviewed by the Connecticut OCME and deemed not human subjects research by the Yale University Human Investigations Committee.

### Geospatial Data

All residential, injury, and death addresses were geocoded within ArcGIS (ESRI, Redlands, CA). Unmatched addresses were then reviewed manually and geocoded using ArcGIS or the R package *tidygeocoder*^17^. In cases where decedents were listed as homeless or where no address was recorded, these cases were not geocoded. Decedents with residential addresses outside Connecticut were excluded. Injury addresses were used as primary locales for the space-time model and mapping the spatial trends. If injury addresses were unknown, residential addresses were used instead. If both addresses were unknown, death addresses were used. The geocoded overdose deaths were then assigned to one of the 169 corresponding cities or towns in Connecticut.

### Covariates

We used demographic information available for residential population at the city/town level in the Connecticut from the American Community Survey (ACS).^18^ We linked these with the city/town-level ACS 5-year estimates from 2015-2019 data and 2010-2014 data to the opioid-detected overdose death records in 2015-2019 and in 2009-2014, respectively. The covariates were the total population size, proportion of the population by demographic covariates (i.e., age groups, sex, race, ethnicity and education level), proportion foreign born, the proportion with home ownership, median household income, and poverty rate.

### Statistical Analyses

We first described demographic information of the decedents with opioid-detected fatal overdoses in Connecticut, stratified by fentanyl-detected overdose deaths and non-fentanyl overdose deaths. Then we examined spatiotemporal trends in counts of opioid-detected overdose deaths within towns in Connecticut. We modelled the count of opioid-detected overdose deaths *n*_*it*_ in town *i* during year *t* as independently and identically Poisson distributed variables with mean *λ*_*it*_,

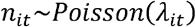

The logarithm of the mean number of opioid-detected overdose deaths (*λ*_*it*_) was then modeled as

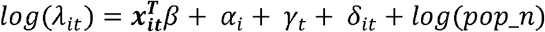

where ***x***_***it***_ is the vector of covariates for town *i* at year *t* (including the time-varying and space-varying variables from the ACS), and *β* is a vector of fixed effect coefficients for ***x***_***it***_. In addition, *α*_*i*_ is the town-level spatial main effect, *γ*_*t*_ is the yearly temporal main effect, and *δ*_*it*_ is the interaction term between space (town level) and time (year). The population size in each city/town was included as an offset *log* (*pop*_*n*) for the Poisson model. The spatial term *α*_*i*_ is a random effect that follows the conditional autoregressive model proposed by Besag, York and Mollie.^19^ The random effect can be further decomposed into two components, an intrinsic conditional autoregressive term that smooths each city/town-level estimate by forming a weighted average with all adjacent jurisdictions, plus a spatially unstructured component that models independent location-specific error and is assumed to be independently, identically, and normally distributed across cities or towns. The temporal trend *γ*_*t*_ is modeled by the sum of two components, a first-order random walk-correlated time component, and a temporally unstructured component that models independent year-specific error and is independently, identically, and normally distributed across calendar years. The space-time interaction term *δ*_*it*_, is modelled as an independent noise term for each city/town and time period and allows for temporal trends in a given city/town to deviate from the overall spatial and temporal trends given by *α*_*i*_ and *γ*_*t*_, so that local patterns can emerge across time and space. Gamma priors were assigned to the precision hyperparameters in our models. Details of the model specification are described in Appendix S1. It should be noted that here we aimed to model the mean count of opioid-detected overdose deaths in each town during each year, and therefore we directly included *λ*_*it*_ in the Poisson model rather than decompose *λ*_*it*_ into expected number and relative risk. In addition, given that there were a considerable number of towns with no opioid overdose deaths especially in the earlier years during the study period, we implemented the zero-inflated Poisson model to account for excess zeroes. The model comparison of the Poisson and zero-inflated Poisson models are shown in Appendix S2. We also conducted a sensitivity analysis that used penalized complexity (PC) priors^20,21^ to the precision hyperparameters instead of gamma priors (see Appendix S3).

To investigate the conditional probability of fentanyl detection given that a fatal opioid-involved overdose occurred, we modelled the probability of an opioid overdose death being fentanyl-detected at the city/town level, using a Bayesian space-time binomial model.^22^ Unlike the Poisson regression directly modeling the count data, the binomial model represents the probability of fentanyl detection given an opioid-detected death. This can tell us whether the conditional risk of fentanyl detection, given an opioid fatal overdose, varies over space and time. Specifically, we considered a binomial model for the number of fentanyl-detected overdose deaths conditional on the total number of opioid overdose deaths in the town areas. Let *p*_*it*_ be the probability of an opioid overdose death being fentanyl-detected in town *i* during year *t*. We assumed that the number of fentanyl-detected overdose deaths *y*_*it*_ in town *i* during year *t* is distributed as

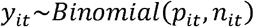

and the corresponding likelihood is

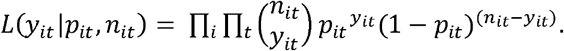

A logistic regression was used to model the probability *p*_*it*_, as

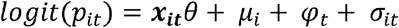

where, similar to model for opioid-detected overdose deaths, ***x***_***it***_ is the vector of covariates for city/town *i* at year *t* (including the time-varying and space-varying variables from the ACS), and *θ* is a vector of fixed effect coefficients for ***x***_***it***_. In addition, *μ*_*i*_ in the model is the city/town-level spatial main effect, *φ*_*t*_ is the yearly temporal main effect, and *σ*_*it*_ is the interaction term between space (city/town level) and time (year). In addition, we conducted three sensitivity analyses to assess the robustness of our results: 1) as an alternative, we constructed a Poisson model for counts of fentanyl-detected overdose deaths among opioid overdose deaths; 2) we restricted the analytical sample to adult (aged ≥18) opioid-detected overdose deaths; 3) we used PC priors to the precision hyperparameters instead of gamma priors.

Finally, we used likelihood ratio tests to examine whether the space-time interaction terms were significantly different from zero in all models. When this term is not significantly different from zero, it suggests that the geographic pattern of fentanyl-detected overdose deaths does not vary significantly over the study period.

To estimate Bayesian model parameters, we employed integrated nested Laplace approximations (INLAs) which approximate the full posterior distribution and are a computationally efficient alternative to Markov Chain Monte Carlo.^23^ Briefly, under certain models (e.g., latent Gaussian models), INLA approximates the marginal posterior distribution of estimated parameters using a multivariate normal distribution.^23,24^ We used the *R-INLA* package for model fitting and estimation.^25^ Model comparison was performed, and details can be found in Appendix S2. All analyses were performed using R Statistical Software (version 4.0.2; R Foundation for Statistical Computing, Vienna, Austria).

## RESULTS

A total of 6,632 opioid-detected overdose deaths were identified by the Connecticut OCME between 2009 and 2019, after excluding 27 deaths with residential addresses outside Connecticut. Among these, 3,234 (49%) were fentanyl-detected, and 3,398 (51%) were non-fentanyl-detected fatalities. The characteristics of these overdose deaths are described in Table 1. People who died of fentanyl-detected overdose deaths were more likely to be male, Black, Hispanic, or involve at least one of the following substances: cocaine, xylazine, gabapentin or mitragynine, compared with those who died of non-fentanyl-detected overdose. The characteristics of overall opioid-detected overdose deaths as well as fentanyl-detected overdose deaths stratified by calendar year are shown in Appendix S4.

**Table 1.**
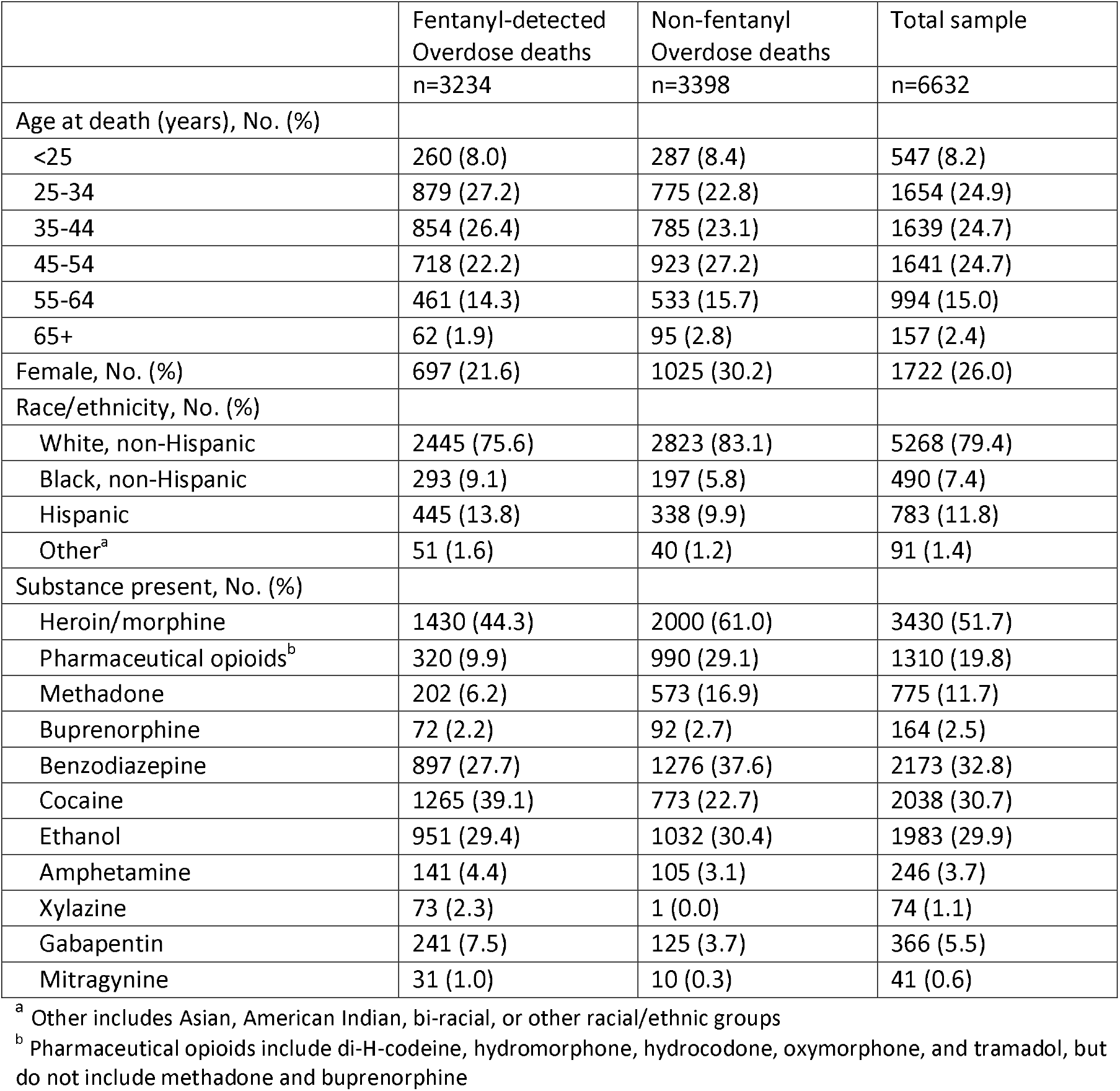
Characteristics of fentanyl-detected overdose deaths among Connecticut residents, 2009-2019

Figure 1 shows the observed temporal trend of fentanyl-detected overdose deaths and non-fentanyl overdose deaths in Connecticut during the study period 2009-2019. Fentanyl-detected overdose deaths increased significantly since 2014. In 2019, 977 (86%) of the 1,138 opioid-detected overdose deaths were fentanyl-detected, compared to 15 (6%) of 266 opioid-detected overdose deaths in 2009 and 36 (8%) of 429 opioid-detected overdose deaths in 2013. The yearly geographic patterns of observed proportion being fentanyl-detected among opioid-detected overdose deaths at the town level are shown in Appendix S5.

**Figure 1.**
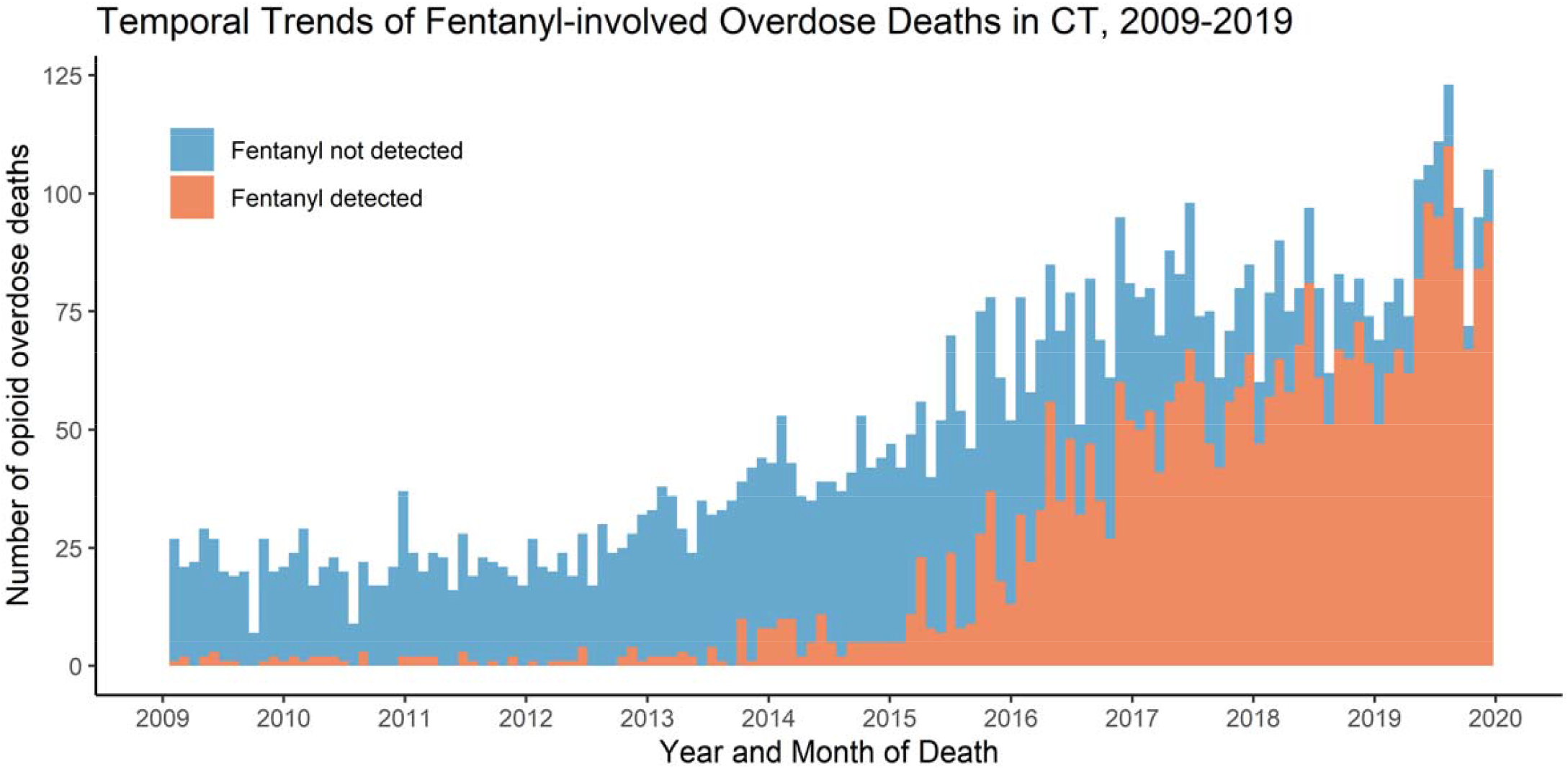
Monthly temporal trend fentanyl-detected overdose deaths among Connecticut residents, 2009-2019. This stacked figure shows, as the number of overdose deaths increased, the share of overdose deaths involving fentanyl increased as well.

Model comparison of the Poisson and zero-inflated Poisson models supported the choice of Poisson model (see Appendix S2). Posterior distributions of estimated parameters for spatial (*α*_*i*_ and temporal (*γ*_*t*_ random effects (while holding other terms constant) of Bayesian space-time Poisson model for overall opioid-detected overdose deaths are shown in Figure 2. Some towns – Torrington and Southington – were at heightened risk of opioid-detected overdose deaths. The temporal trends showed the risk of opioid-detected overdose deaths among residents in Connecticut increased substantially from 2009 to 2019, although there was a slight decrease in 2018.

**Figure 2:**
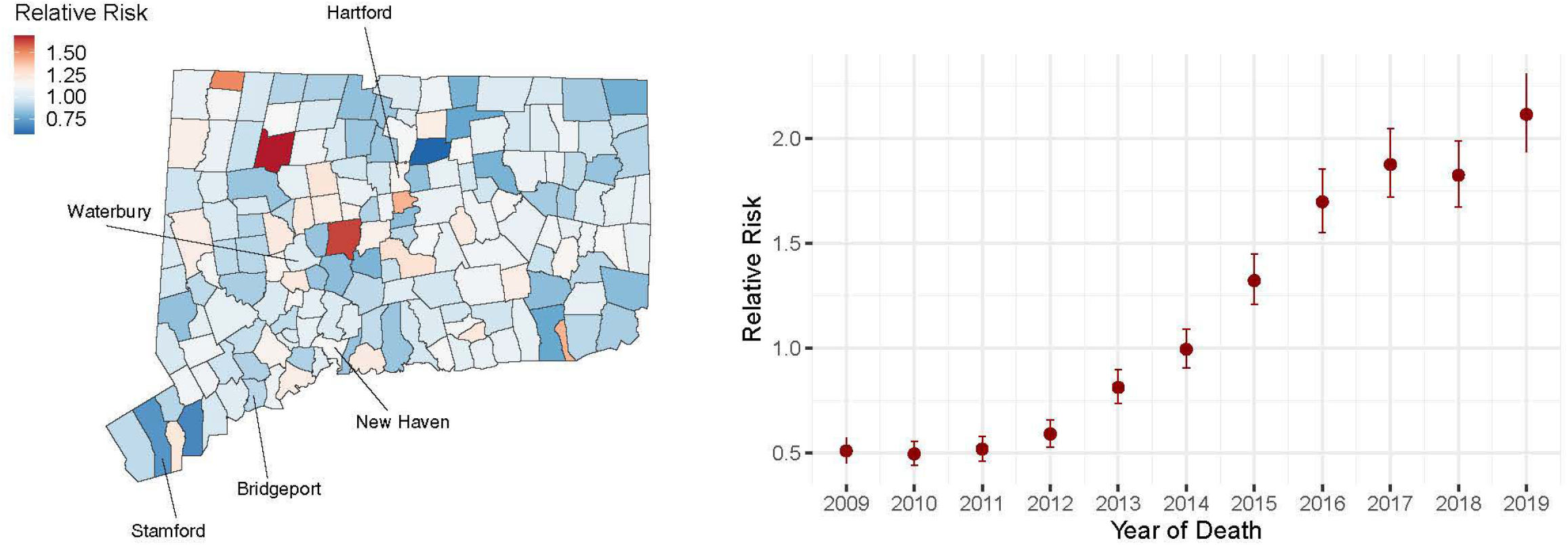
Posterior distributions of estimated parameters for spatial (*α*_*i*_) and temporal (*γ*_*t*_) random effects (relative risk) from the Bayesian space-time Poisson model for overall opioid-detected overdose deaths. The left panel depicts the spatial patterns of overall opioid-detected overdose deaths when holding temporal terms and other covariates constant. The right panel depicts the temporal patterns of overall opioid-detected overdose deaths when holding spatial terms and other covariates constant. The random effects were exponentiated. Larger values of relative risk indicate the higher risk of opioid-detected overdose deaths at town level in Connecticut. Note: link to the Connecticut Towns Index Map (https://portal.ct.gov/-/media/DEEP/gis/Resources/IndexTownspdf.pdf)

Based on the fitted Bayesian space-time binomial model for the proportion of fentanyl-detected overdose deaths, posterior distributions of the fixed-effect coefficients (*θ*) of covariates are summarized in Figure 3. Increased proportion of residents aged 55-64 in a city/town was associated with decreased probability being fentanyl-detected, given an opioid-detected overdose death in that town, with an odds ratio (OR) of 0.92 (95% credible interval [CrI], 0.86, 0.99). In addition, the proportion in poverty was negatively associated with the probability of a fatal overdose being fentanyl-detected (OR=0.95 [95% CrI, 0.91, 0.99]).

**Figure 3.**
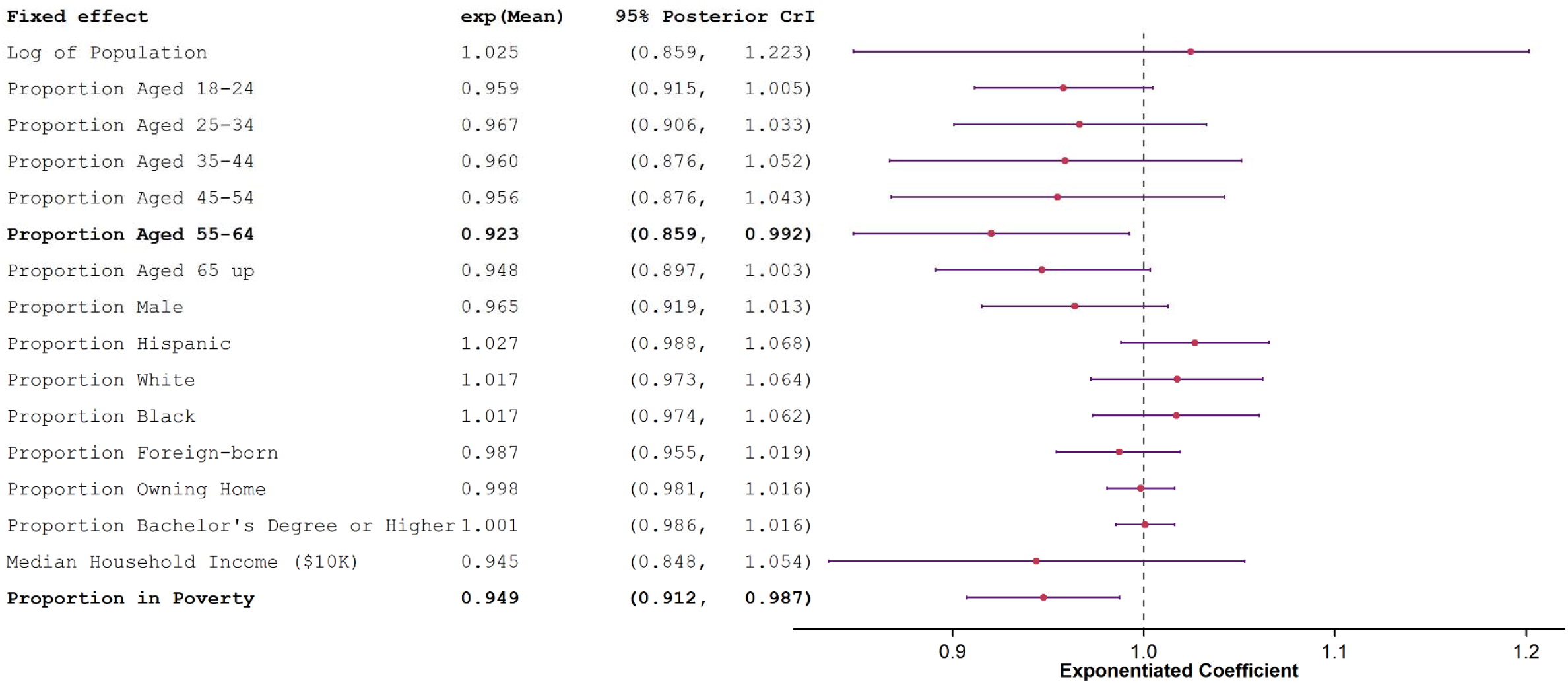
Summary of the posterior means and 95% credible intervals (CrIs) of the fixed effect coefficients of covariates. The regression coefficients are exponentiated to represent exponentiated coefficients from binomial logistic regression.

Posterior distributions of estimated parameters for spatial (*μ*_*i*_) and temporal (*φ*_*t*_) random effects (while holding other terms constant) are summarized in Figure 4, along with geographic patterns across towns. Some areas had increased probability of fentanyl involvement, and the spatial modeling (holding temporal terms constant) suggests that even after adjusting for the city/town level demographic covariates, the northeastern region of Connecticut had a higher probability of fentanyl-detected deaths, with Hartford, East Hartford and Manchester at the highest risk. In contrast, New Haven and its surrounding towns and the southwestern Connecticut—primarily Greenwich—had a lower probability of fentanyl-detected deaths. The temporal trends in Figure 4 describe main temporal trends similar to those observed in Figure 1, that is, that the probability of an opioid overdose death being fentanyl-detected increased monotonically since 2014.

**Figure 4:**
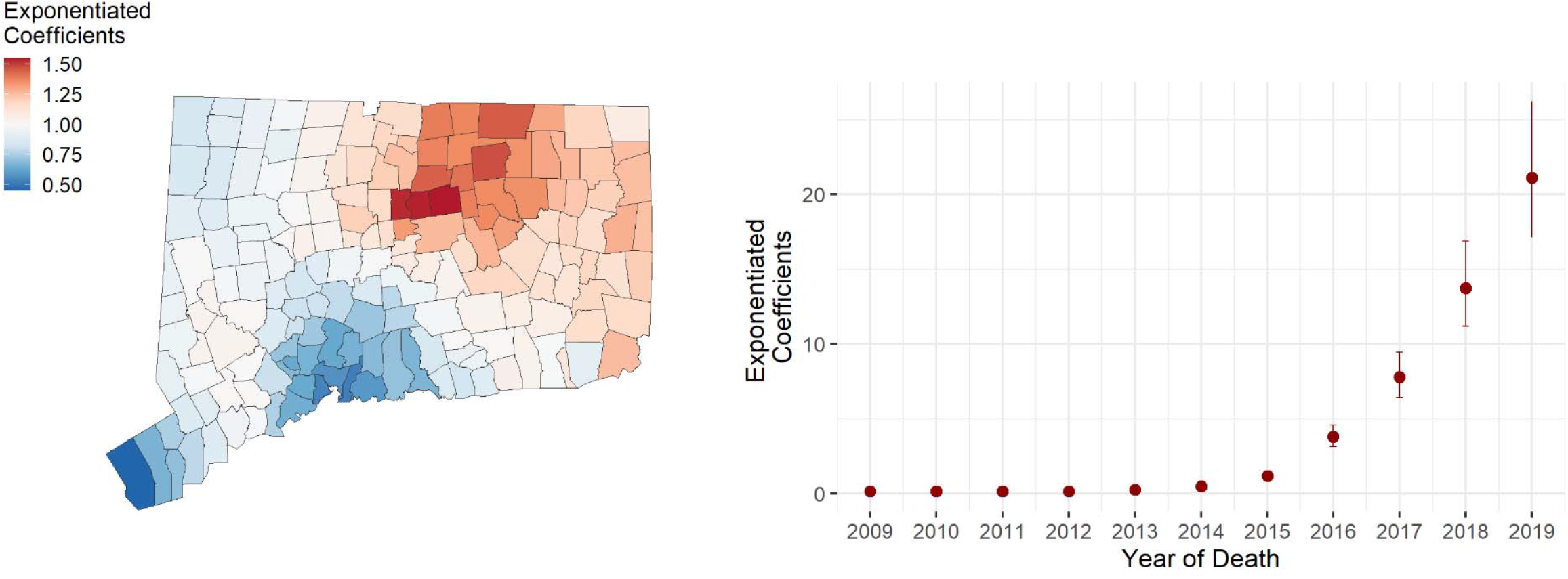
Posterior distributions of estimated parameters for spatial (*μ*_*i*_) and temporal (*φ*_*t*_) random effects from the Bayesian space-time binomial model for fentanyl-detected overdose deaths. The left panel depicts the spatial patterns of fentanyl-detected overdose deaths when holding temporal terms and other covariates constant. The right panel depicts the temporal patterns of fentanyl-detected overdose deaths when holding spatial terms and other covariates constant. The random effects were exponentiated. Larger values of exponentiated coefficients indicate the higher probability being fentanyl detected given an opioid overdose death at town level in Connecticut. Note: link to the Connecticut Towns Index Map (https://portal.ct.gov/-/media/DEEP/gis/Resources/IndexTownspdf.pdf)

The city/town-level predicted probabilities of being a fentanyl-detected overdose death from the Bayesian space-time binomial model are shown in Figure 5. A higher probability being fentanyl-detected, given an opioid overdose death, was first found in several towns in the northeastern region of Connecticut during 2009-2013. Beginning in 2014, the fentanyl “hotspots” started to spread across Connecticut.

**Figure 5.**
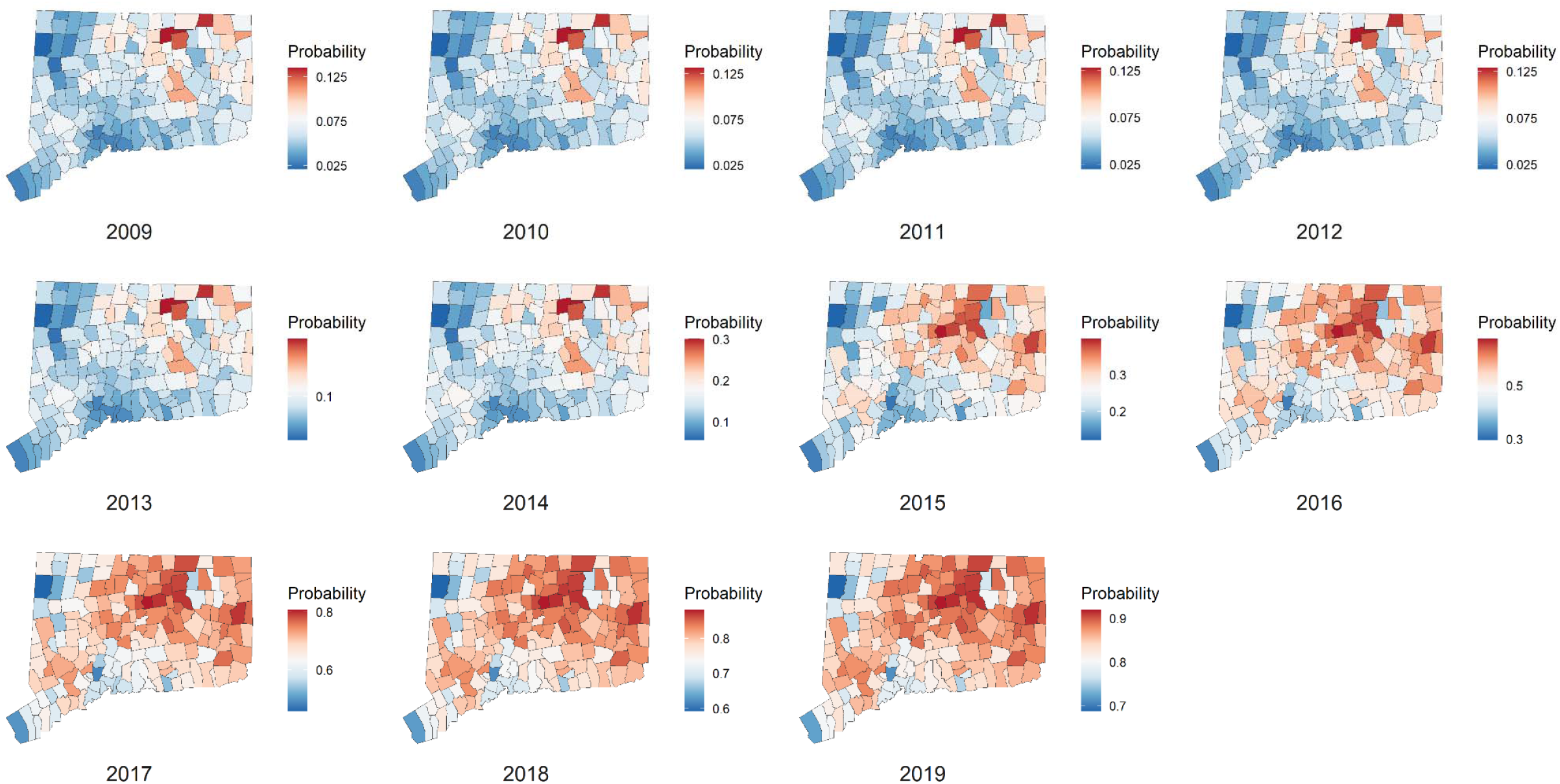
The predicted probability of an overdose death being fentanyl-detected from the Bayesian space-time binomial model, stratified by town and calendar year, Connecticut. Color scales differ by year to show the distribution within each year. Note: link to the Connecticut Towns Index Map (https://portal.ct.gov/-/media/DEEP/gis/Resources/IndexTownspdf.pdf)

A likelihood ratio test shows that the space-time interaction term of the Bayesian space-time binomial model for the proportion of fentanyl-detected opioid overdose deaths was not significantly different from zero, suggesting the geographic patterns of fentanyl-detected overdose deaths gradually overtook the preceding non-fentanyl opioid overdose deaths.

The results of the sensitivity analyses 1) using Poisson model for fentanyl-detected overdose deaths in Connecticut, 2) restricting to adult opioid-detected overdose deaths, or 3) using PC priors for prior distribution of hyperparameters were similar to those using a binomial logistic model and are described in Appendix S6.

## DISCUSSION

In the present study we examined the town-level geographic patterns and yearly temporal trends of fentanyl-detected overdose deaths among Connecticut residents during 2009-2019 and evaluated the relationship of fentanyl-detected overdose deaths with overall trends of opioid-detected overdose deaths. To our knowledge, this is the first study to map the spatiotemporal distributions of fentanyl-detected overdose deaths in Connecticut. We identified regions of Connecticut, particularly in the northeastern part of the state, as having relatively high probability of an opioid overdose death being fentanyl-detected. The temporal trends show the fentanyl-detected overdose deaths in Connecticut increased significantly since 2014. We also found that the geographic pattern of fentanyl-detected overdose deaths was relatively constant over time and gradually replaced the preceding wave of opioid overdose deaths.

Our findings show that, compared with non-fentanyl overdose deaths, people who died of fentanyl-detected overdose are more likely to be Black or Hispanic in Connecticut, suggesting that these groups are disproportionately affected by the most recent waves of opioid crisis. These results are consistent with the findings from previous studies that examined opioid-detected overdose deaths in other regions and across the US.^26–28^ More research is warranted to address the social determinants of fentanyl-detected overdose deaths among Black and Hispanic communities, and locally informed harm reduction efforts and services should be targeted to these populations.

Our results indicated that the northeastern region of Connecticut had higher probability of an opioid overdose death being fentanyl-detected whereas New Haven, its surrounding towns, and the southwestern Connecticut had a lower probability. This suggests that the Connecticut supply of fentanyl and its analogues may have originally entered Connecticut from the north, gradually diffused from the northeastern to southwestern Connecticut, and most recently has dispersed across the state. Our findings also suggest that the fentanyl-detected overdose deaths might have simply replaced the preceding waves of opioid overdose deaths (e.g., heroin), rather than creating new overdose risk following distinct geographic patterns. These findings were in line with the facts that fentanyl and its analogues entered into illicit drug supply as an “adulterant” in powder heroin and counterfeit pills in Northeast.^29^ The growing presence and strong potency of fentanyl and its analogues then led to the significant increase in opioid overdose deaths across Connecticut in recent years.

Further studies are warranted to explore the potential factors (e.g., supply-side aspects, neighborhood characteristics, and availability and accessibility of health services and naloxone distribution) that result in geographical variability in fentanyl-detected overdose mortality. For example, a recent study in Canada showed the rural areas experienced a disproportionately high risk of fatal overdoses from illicit drug poisoning compared with urban areas.^30^ Such rural-urban differences may be due to limited access to harm reduction services and rising local drug toxicity. Our study provides insight into the distinctive geographic patterns in fentanyl mortality in Connecticut. These findings can inform targeted overdose surveillance in specific regions of the state and guide more efficient deployment of harm reduction services and opioid treatment programs in Connecticut.

Our study is subject to several limitations. First, although we adjusted for city/town-level demographic covariates in our Bayesian space-time models, it is possible that this ecological analysis obscures individual-level relationships between time, place, and overdose risk. We described the geographic patterns and temporal trends of fentanyl-detected overdose deaths at the city/town level only and could not examine in detail the potential social determinants for an opioid overdose death being fentanyl-detected. Second, we primarily used injury addresses for the geographic locales and used residential or death addresses only when injury address was missing, which may lead to geographical misclassification. However, most deaths occurred within person’s own residence so that the injury and residential addresses were the same. Moreover, death addresses were often in the hospital near the injury place. Therefore, we expect that the influence of geographic misclassification on our study findings was minimal. Lastly, while we investigated the spatiotemporal trends of fentanyl-detected overdose deaths, we did not explore the trends of overdose deaths that involved both fentanyl and other substances such as methamphetamine. Further studies are needed to investigate the spatiotemporal trends of polysubstance use with fentanyl in Connecticut.

In conclusion, given the high probability of an overdose death being fentanyl-detected, more research should be devoted to exploring the social determinants and supply-side drivers of fentanyl-involved overdose deaths in order to better inform future harm reduction services and policies to halt the current opioid crisis.

## Data Availability

All data produced in the present study are available upon reasonable request to the authors

## Acknowledgements

We are grateful to Dr. James R. Gill and his staff at the Connecticut Office of the Chief Medical Examiner for providing access to data. In addition, we thank Joshua L. Warren, Thomas A. Thornhill, Julia Dennett, and A Ram for helpful comments.

## Appendix

### Appendix S1

#### Priors on random effects in Bayesian space-time Poisson model for overall opioid-detected overdose deaths

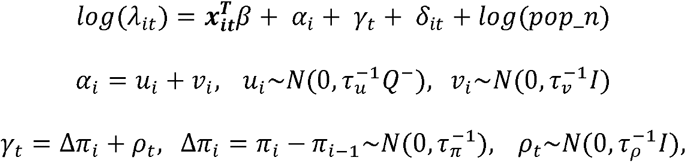

where 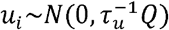 represents the spatial structured random effect and is modeled under the class of intrinsic Gaussian Markov random fields models. *Q* denotes the precision matrix (neighboring matrix), and *Q*^−^ is the generalized inverse of the matrix *Q*. The marginal variances are 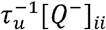, which are dependent on the matrix *Q*. 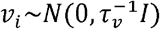 is the spatial unstructured random effect and 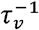 is the marginal variance. Gamma priors with small rate parameters are commonly assigned to *τ*_*u*_ and *τ*_*v*_. Here, Gamma(1, 0.0005) is considered.

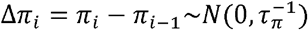 is first order random walk temporal random effect defined as a random step at each point in time (∆*π*_*i*_). All random steps are independent and identically distributed. 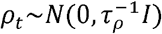 is the temporal unstructured random effect and 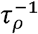 is the marginal variance. Gamma priors with small rate parameters are commonly assigned to *τ*_*π*_ and *τ*_*ρ*_. Here, Gamma(1, 0.00005) is considered.

### Appendix S2: Model Comparison

#### Appendix S2.1: Model Comparison for overall opioid-detected overdose deaths

For overall opioid-detected overdose deaths, Poisson models were used to estimate the mean number of opioid-detected overdose deaths in each town at each year from 2009-2019. Concerning some towns with 0 opioid-detected overdose deaths, zero-inflated Poisson models were also examined. Deviance information criterion (DIC) and Watanabe-Akaike information criterion were used to compare Poisson model and zero-inflated Poisson model.

**Appendix S2. Table 1:** Model Comparison for overall opioid-detected overdose deaths

| Model | DIC | WAIC |
| --- | --- | --- |
| Poisson (in the main text) | 5726.50 | 5733.06 |
| Zero-inflated Poisson | 6525.00 | 6528.64 |

Based on DIC and WAIC, Poisson model is selected.

#### Appendix S2.2: Model Comparison for fentanyl-detected overdose deaths

For fentanyl-detected overdose death among all opioid-detected overdose deaths, binomial model and logistic regression were used to estimate the probability being fentanyl-detected given an opioid overdose death in each town at each year from 2009-2019. Several parsimonious models, in addition to the logistic model mentioned in the main text were examined. DIC and WAIC were used for model comparison.

**Appendix S2. Table 2:**
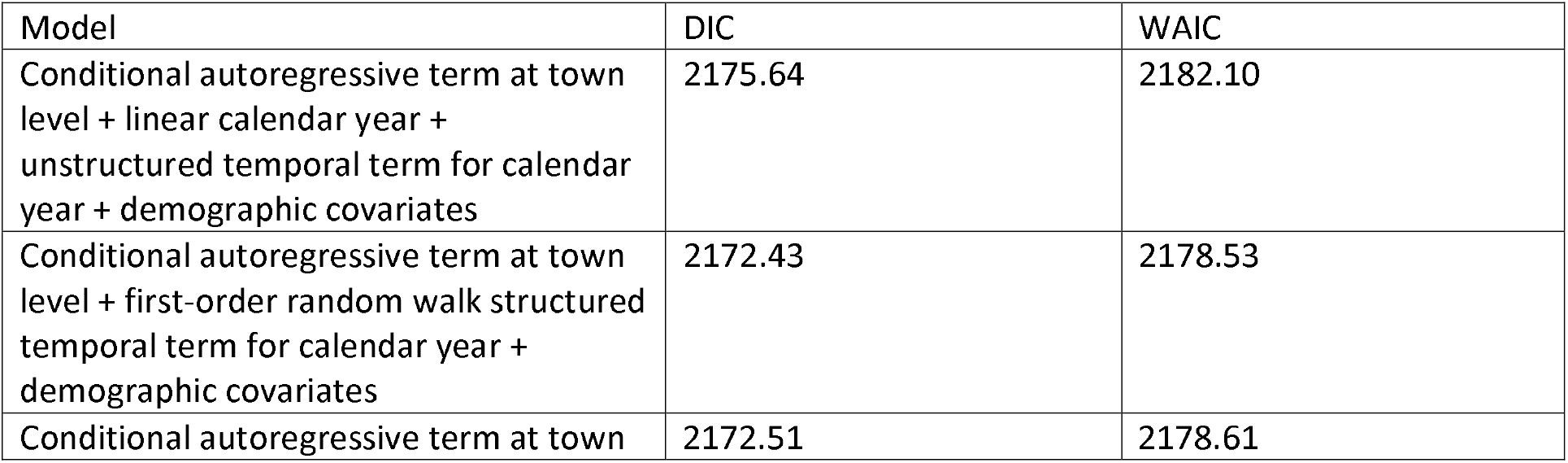
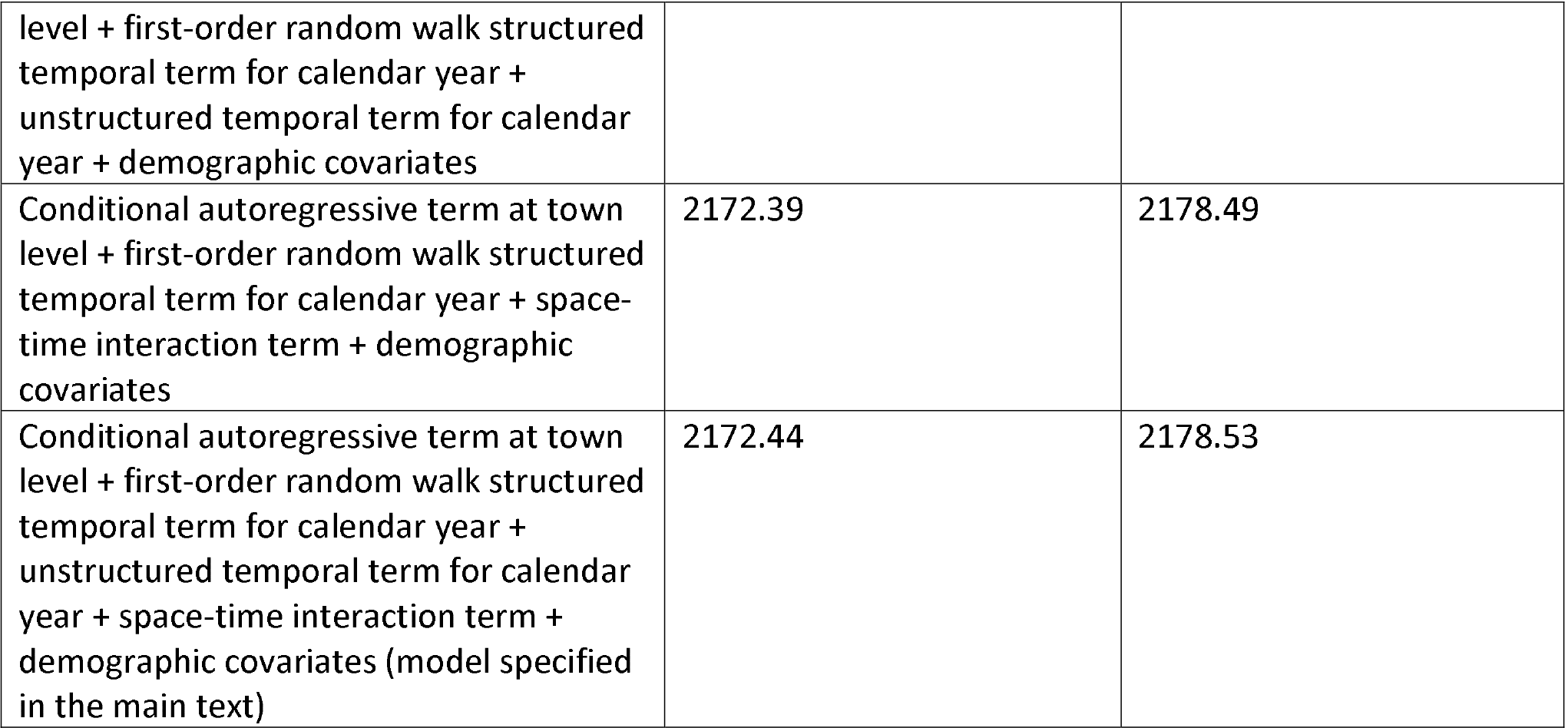
Model Comparison for fentanyl-detected overdose deaths

Based on DIC and WAIC, the binomial model including conditional autoregressive term at town level, first-order random walk structured temporal term for calendar year, space-time interaction term, and demographic covariates, without unstructured temporal term, was selected.

### Appendix S3

#### Sensitivity analysis using penalized complexity (PC) priors for Bayesian space-time Poisson model for overall opioid-detected overdose deaths

In this sensitivity analysis, instead of using gamma priors for precision hyperparameters in Bayesian space-time Poisson model for overall opioid-detected overdose deaths, we employed penalized complexity (PC)^1^ priors for the prior distribution of hyperparameters. The hyperprior distributions are chosen with the PC framework.^2^ We let the precision parameters *τ*_*u*_, *τ*_*v*_, *τ*_*π*_ and *τ*_*ρ*_ (as described in Appendix S1) ∼*PC*(0.2/0.31, 0.01), which corresponds to 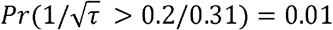, leading to the prior standard deviation of the marginal log relative risk being 0.2. The results of this sensitivity analysis were shown in Appendix S3 Figure 1.

---

**Appendix S3. Figure 1:**
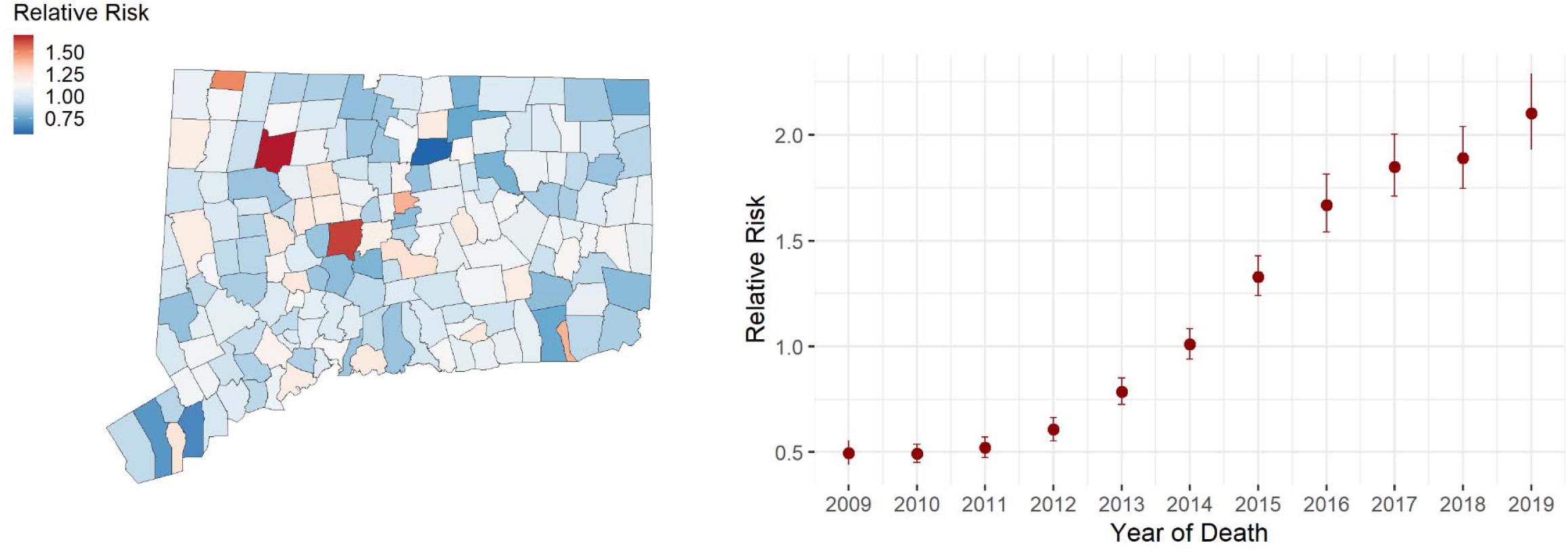
Posterior distributions of estimated parameters for spatial (*α*_*i*_) and temporal (*γ*_*t*_) random effects (relative risk) from the Bayesian space-time Poisson model for overall opioid-detected overdose deaths using penalized complexity priors for hyperparameters. The left panel depicts the spatial patterns of overall opioid-detected overdose deaths when holding temporal terms and other covariates constant. The right panel depicts the temporal patterns of overall opioid-detected overdose deaths when holding spatial terms and other covariates constant. The random effects were exponentiated. Larger values of relative risk indicate the higher risk of opioid-detected overdose deaths at town level in Connecticut. Note: link to the Connecticut Towns Index Map (https://portal.ct.gov/-/media/DEEP/gis/Resources/IndexTownspdf.pdf)

## Appendix S4

**Appendix S4. Table 1.** Characteristics of opioid-detected overdose deaths among Connecticut residents by year, 2009-2019.

| Year | 2009 | 2010 | 2011 | 2012 | 2013 | 2014 | 2015 | 2016 | 2017 | 2018 | 2019 |
| --- | --- | --- | --- | --- | --- | --- | --- | --- | --- | --- | --- |
|  | n=266 | n=254 | n=268 | n=298 | n=429 | n=510 | n=678 | n=883 | n=960 | n=949 | n=1138 |
| Age at death (years), No. (%) |  |  |  |  |  |  |  |  |  |  |  |
| <25 | 28 (10.5) | 26 (10.2) | 39 (14.6) | 39 (13.1) | 48 (11.2) | 31 (6.1) | 60 (8.8) | 65 (7.4) | 79 (8.2) | 68 (7.2) | 64 (5.6) |
| 25-34 | 62 (23.3) | 45 (17.7) | 75 (28.1) | 74 (24.8) | 106 (24.7) | 131 (25.7) | 167 (24.6) | 233 (26.4) | 264 (27.5) | 226 (23.8) | 271 (23.8) |
| 35-44 | 81 (30.5) | 72 (28.3) | 58 (21.7) | 60 (20.1) | 95 (22.1) | 132 (25.9) | 146 (21.5) | 200 (22.7) | 239 (24.9) | 248 (26.1) | 308 (27.1) |
| 45-54 | 64 (24.1) | 91 (35.8) | 64 (24.0) | 87 (29.2) | 113 (26.3) | 142 (27.8) | 173 (25.5) | 225 (25.5) | 213 (22.2) | 229 (24.1) | 240 (21.1) |
| 55-64 | 29 (10.9) | 20 (7.9) | 30 (11.2) | 30 (10.1) | 62 (14.5) | 59 (11.6) | 110 (16.2) | 144 (16.3) | 148 (15.4) | 148 (15.6) | 214 (18.8) |
| ≥65 | 2 (0.8) | 0 (0.0) | 1 (0.4) | 8 (2.7) | 5 (1.2) | 15 (2.9) | 22 (3.2) | 16 (1.8) | 17 (1.8) | 30 (3.2) | 41 (3.6) |
| Female, No. (%) | 83 (31.2) | 79 (31.1) | 76 (28.5) | 80 (26.8) | 127 (29.6) | 154 (30.2) | 178 (26.3) | 228 (25.8) | 232 (24.2) | 222 (23.4) | 263 (23.1) |
| Race/ethnicity, No. (%) |  |  |  |  |  |  |  |  |  |  |  |
| White, non-Hispanic | 228 (85.7) | 214 (84.3) | 236 (88.4) | 245 (82.2) | 357 (83.2) | 430 (84.3) | 562 (82.9) | 696 (78.8) | 764 (79.6) | 710 (74.8) | 826 (72.6) |
| Black, non-Hispanic | 12 (4.5) | 13 (5.1) | 9 (3.4) | 20 (6.7) | 26 (6.1) | 21 (4.1) | 36 (5.3) | 64 (7.2) | 72 (7.5) | 94 (9.9) | 123 (10.8) |
| Hispanic | 25 (9.4) | 26 (10.2) | 20 (7.5) | 28 (9.4) | 42 (9.8) | 56 (11.0) | 74 (10.9) | 102 (11.6) | 111 (11.6) | 127 (13.4) | 172 (15.1) |
| Other<sup>a</sup> | 1 (0.4) | 1 (0.4) | 2 (0.7) | 5 (1.7) | 4 (0.9) | 3 (0.6) | 6 (0.9) | 21 (2.4) | 13 (1.4) | 18 (1.9) | 17 (1.5) |
| Substance present, No. (%) |  |  |  |  |  |  |  |  |  |  |  |
| Fentanyl/fentanyl analogs | 15 (5.6) | 14 (5.5) | 14 (5.2) | 15 (5.0) | 36 (8.4) | 75 (14.7) | 183 (27.0) | 472 (53.5) | 670 (69.8) | 763 (80.4) | 977 (85.9) |
| Heroin/morphine | 146 (54.9) | 124 (48.8) | 144 (53.9) | 181 (60.7) | 278 (64.8) | 335 (65.7) | 432 (63.7) | 522 (59.1) | 463 (48.2) | 399 (42.0) | 406 (35.7) |
| Pharmaceutical opioids<sup>b</sup> | 96 (36.1) | 96 (37.8) | 80 (30.0) | 81 (27.2) | 108 (25.2) | 142 (27.8) | 162 (23.9) | 155 (17.6) | 149 (15.5) | 107 (11.3) | 134 (11.8) |
| Methadone | 66 (24.8) | 58 (22.8) | 55 (20.6) | 37 (12.4) | 48 (11.2) | 54 (10.6) | 81 (11.9) | 96 (10.9) | 100 (10.4) | 84 (8.9) | 96 (8.4) |
| Buprenorphine | 0 (0.0) | 0 (0.0) | 0 (0.0) | 0 (0.0) | 7 (1.6) | 9 (1.8) | 21 (3.1) | 30 (3.4) | 25 (2.6) | 29 (3.1) | 43 (3.8) |
| Benzodiazepine | 62 (23.3) | 84 (33.1) | 93 (34.8) | 48 (16.1) | 131 (30.5) | 225 (44.1) | 285 (42.0) | 304 (34.4) | 345 (35.9) | 284 (29.9) | 312 (27.4) |
| Cocaine | 38 (14.3) | 34 (13.4) | 64 (24.0) | 67 (22.5) | 113 (26.3) | 118 (23.1) | 162 (23.9) | 263 (29.8) | 333 (34.7) | 365 (38.5) | 481 (42.3) |
| Ethanol | 69 (25.9) | 76 (29.9) | 87 (32.6) | 60 (20.1) | 114 (26.6) | 155 (30.4) | 188 (27.7) | 274 (31.0) | 321 (33.4) | 274 (28.9) | 365 (32.1) |
| Amphetamine | 1 (0.4) | 1 (0.4) | 3 (1.1) | 5 (1.7) | 5 (1.2) | 22 (4.3) | 35 (5.2) | 29 (3.3) | 35 (3.6) | 48 (5.1) | 62 (5.4) |
| Xylazine | 0 (0.0) | 0 (0.0) | 0 (0.0) | 0 (0.0) | 0 (0.0) | 0 (0.0) | 0 (0.0) | 0 (0.0) | 0 (0.0) | 0 (0.0) | 74 (6.5) |
| Gabapentin | 0 (0.0) | 0 (0.0) | 1 (0.4) | 0 (0.0) | 0 (0.0) | 0 (0.0) | 2 (0.3) | 50 (5.7) | 85 (8.9) | 77 (8.1) | 151 (13.3) |
| Mitragynine | 0 (0.0) | 0 (0.0) | 0 (0.0) | 0 (0.0) | 0 (0.0) | 1 (0.2) | 0 (0.0) | 8 (0.9) | 3 (0.3) | 7 (0.7) | 22 (1.9) |

<sup>a</sup> Other includes Asian, American Indian, bi-racial, or other racial/ethnic groups
<sup>b</sup> Pharmaceutical opioids include di-H-codeine, hydromorphone, hydrocodone, oxycodone, and tramadol, but do not include methadone and buprenorphine

**Appendix S4. Table 2.** Characteristics of fentanyl-detected overdose deaths among Connecticut residents by year, 2009-2019.

| Year | 2009 | 2010 | 2011 | 2012 | 2013 | 2014 | 2015 | 2016 | 2017 | 2018 | 2019 |
| --- | --- | --- | --- | --- | --- | --- | --- | --- | --- | --- | --- |
|  | n=266 | n=254 | n=268 | n=298 | n=429 | n=510 | n=678 | n=883 | n=960 | n=949 | n=1138 |
| Age at death (years), No. (%) |  |  |  |  |  |  |  |  |  |  |  |
| <25 | 2 (13.3) | 1 (7.1) | 4 (28.6) | 1 (6.7) | 2 (5.6) | 7 (9.3) | 19 (10.4) | 35 (7.4) | 64 (9.6) | 63 (8.3) | 62 (6.3) |
| 25-34 | 3 (20.0) | 4 (28.6) | 4 (28.6) | 3 (20.0) | 6 (16.7) | 21 (28.0) | 50 (27.3) | 136 (28.8) | 209 (31.2) | 195 (25.6) | 248 (25.4) |
| 35-44 | 2 (13.3) | 2 (14.3) | 3 (21.4) | 1 (6.7) | 8 (22.2) | 25 (33.3) | 43 (23.5) | 122 (25.8) | 175 (26.1) | 201 (26.3) | 272 (27.8) |
| 45-54 | 4 (26.7) | 6 (42.9) | 1 (7.1) | 6 (40.0) | 12 (33.3) | 16 (21.3) | 47 (25.7) | 111 (23.5) | 138 (20.6) | 177 (23.2) | 200 (20.5) |
| 55-64 | 4 (26.7) | 1 (7.1) | 2 (14.3) | 3 (20.0) | 8 (22.2) | 4 (5.3) | 22 (12.0) | 61 (12.9) | 79 (11.8) | 110 (14.4) | 167 (17.1) |
| ≥65 | 0 (0.0) | 0 (0.0) | 0 (0.0) | 1 (6.7) | 0 (0.0) | 2 (2.7) | 2 (1.1) | 7 (1.5) | 5 (0.7) | 17 (2.2) | 28 (2.9) |
| Female, No. (%) | 9 (60.0) | 4 (28.6) | 4 (28.6) | 5 (33.3) | 12 (33.3) | 25 (33.3) | 34 (18.6) | 94 (19.9) | 136 (20.3) | 162 (21.2) | 212 (21.7) |
| Race/ethnicity, No. (%) |  |  |  |  |  |  |  |  |  |  |  |
| White, non-Hispanic | 12 (80.0) | 11 (78.6) | 13 (92.9) | 14 (93.3) | 26 (72.2) | 61 (81.3) | 149 (81.4) | 370 (78.4) | 523 (78.1) | 568 (74.4) | 698 (71.4) |
| Black, non-Hispanic | 1 (6.7) | 1 (7.1) | 0 (0.0) | 0 (0.0) | 4 (11.1) | 4 (5.3) | 9 (4.9) | 33 (7.0) | 55 (8.2) | 79 (10.4) | 107 (11.0) |
| Hispanic | 2 (13.3) | 2 (14.3) | 1 (7.1) | 1 (6.7) | 6 (16.7) | 9 (12.0) | 23 (12.6) | 62 (13.1) | 82 (12.2) | 101 (13.2) | 156 (16.0) |
| Other<sup>a</sup> | 0 (0.0) | 0 (0.0) | 0 (0.0) | 0 (0.0) | 0 (0.0) | 1 (1.3) | 2 (1.1) | 7 (1.5) | 10 (1.5) | 15 (2.0) | 16 (1.6) |
| Substance present, No. (%) |  |  |  |  |  |  |  |  |  |  |  |
| Heroin/morphine | 2 (13.3) | 1 (7.1) | 2 (14.3) | 1 (6.7) | 11 (30.6) | 38 (50.7) | 112 (61.2) | 277 (58.7) | 321 (47.9) | 313 (41.0) | 352 (36.0) |
| Pharmaceutical opioids<sup>b</sup> | 4 (26.7) | 2 (14.3) | 4 (28.6) | 3 (20.0) | 6 (16.7) | 14 (18.7) | 25 (13.7) | 53 (11.2) | 66 (9.9) | 60 (7.9) | 83 (8.5) |
| Methadone | 2 (13.3) | 2 (14.3) | 0 (0.0) | 1 (6.7) | 3 (8.3) | 5 (6.7) | 6 (3.3) | 33 (7.0) | 43 (6.4) | 40 (5.2) | 67 (6.9) |
| Buprenorphine | 0 (0.0) | 0 (0.0) | 0 (0.0) | 0 (0.0) | 0 (0.0) | 1 (1.3) | 6 (3.3) | 12 (2.5) | 10 (1.5) | 16 (2.1) | 27 (2.8) |
| Benzodiazepine | 4 (26.7) | 3 (21.4) | 4 (28.6) | 2 (13.3) | 9 (25.0) | 23 (30.7) | 56 (30.6) | 127 (26.9) | 208 (31.0) | 213 (27.9) | 248 (25.4) |
| Cocaine | 3 (20.0) | 2 (14.3) | 3 (21.4) | 1 (6.7) | 16 (44.4) | 19 (25.3) | 50 (27.3) | 157 (33.3) | 254 (37.9) | 320 (41.9) | 440 (45.0) |
| Ethanol | 1 (6.7) | 4 (28.6) | 3 (21.4) | 0 (0.0) | 9 (25.0) | 14 (18.7) | 47 (25.7) | 140 (29.7) | 214 (31.9) | 215 (28.2) | 304 (31.1) |
| Amphetamine | 0 (0.0) | 0 (0.0) | 0 (0.0) | 0 (0.0) | 0 (0.0) | 2 (2.7) | 10 (5.5) | 13 (2.8) | 19 (2.8) | 41 (5.4) | 56 (5.7) |
| Xylazine | 0 (0.0) | 0 (0.0) | 0 (0.0) | 0 (0.0) | 0 (0.0) | 0 (0.0) | 0 (0.0) | 0 (0.0) | 0 (0.0) | 0 (0.0) | 73 (7.5) |
| Gabapentin | 0 (0.0) | 0 (0.0) | 0 (0.0) | 0 (0.0) | 0 (0.0) | 0 (0.0) | 0 (0.0) | 22 (4.7) | 48 (7.2) | 50 (6.6) | 121 (12.4) |
| Mitragynine | 0 (0.0) | 0 (0.0) | 0 (0.0) | 0 (0.0) | 0 (0.0) | 0 (0.0) | 0 (0.0) | 3 (0.6) | 2 (0.3) | 5 (0.7) | 21 (2.1) |

<sup>a</sup> Other includes Asian, American Indian, bi-racial, or other racial/ethnic groups
<sup>b</sup> Pharmaceutical opioids include di-H-codeine, hydromorphone, hydrocodone, oxycodone, and tramadol, but do not include methadone and buprenorphine

## Appendix S5

**Appendix S5. Figure 1.**
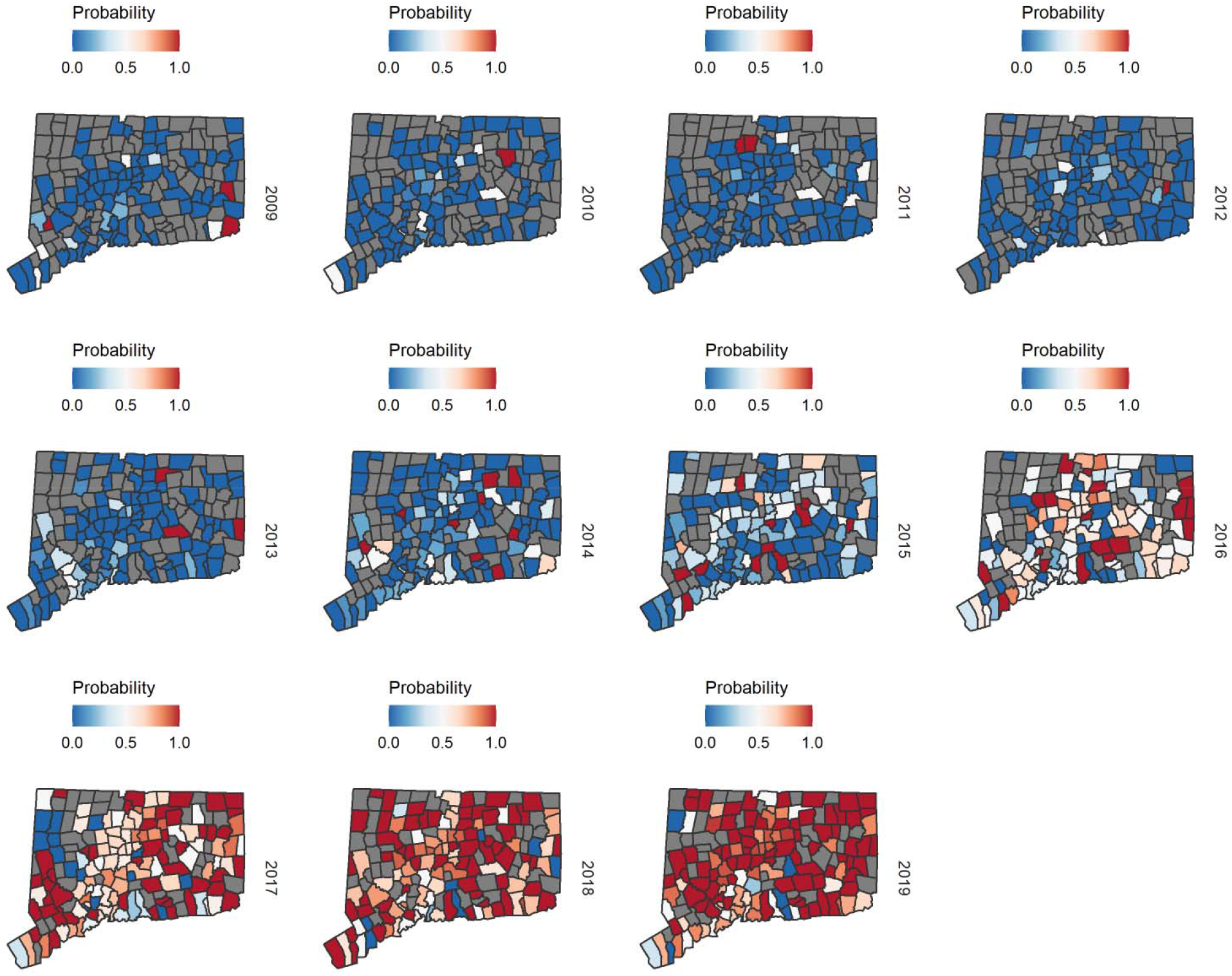
Spatial patterns of observed proportion being fentanyl-detected among opioid overdose deaths at the city/town level, stratified by year, 2009-2019. Towns with grey color represent no opioid-detected deaths. Note: link to the Connecticut Towns Index Map (https://portal.ct.gov/-/media/DEEP/gis/Resources/IndexTownspdf.pdf)

## Appendix S6

### Sensitivity Analysis 1

Poisson model was used as an alternative to binomial model to estimate the risk of fentanyl-detected overdose deaths among opioid-detected overdose deaths at town level in Connecticut, 2009-2019. Specifically, we modelled the number of fentanyl-detected overdose deaths *y*_*it*_ in town *i* during year *t* as independently and identically Poisson distributed variables with mean *µ*_*it*_,

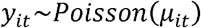

Then the logarithm of the mean number of fentanyl-detected overdose deaths (*µ*_*it*_) is modeled as

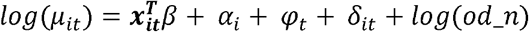

where ***x***_***it***_ is the vector of covariates for town *i* at year *t* (including the time-varying and space-varying variables from ACS as aforementioned), and *β* is a vector of fixed effect coefficients for ***x***_***it***_. In addition, *α*_*i*_ in the model is town-level spatial main effect, *φ*_*t*_ is the yearly temporal main effect, and *δ*_*it*_ is the interaction term between space (town level) and time (year). The number of overall opioid-detected overdose deaths in each town in each year was used as an offset *log*(*od*_*n*).

Specifically, the spatial term *α*_*i*_ is a random effect that follows the conditional autoregressive model proposed by Besag, York and Mollie.^19^ The random effect can be further decomposed into two components, an intrinsic conditional autoregressive term that smooths each town-level estimate by forming a weighted average with all adjacent census tracts, plus spatially unstructured component that models independent location-specific error and is assumed to be independently identically normally distributed across towns. The temporal trend *φ*_*t*_, is modeled by the sum of two components, a first-order random walk-correlated time component, and a temporally unstructured component that models independent year-specific error and is independently identically and normally distributed across calendar years. The space-time interaction term *δ*_*it*_, is modelled as an independent noise term for each town and time period, and allows for temporal trends in a given towns to deviate from the overall spatial and temporal trends given by *α*_*i*_ and *φ*_*t*_, such that local patterns can emerge across time and space. The results were shown in Appendix S6 Figure 1.

**Appendix S6. Figure 1.**
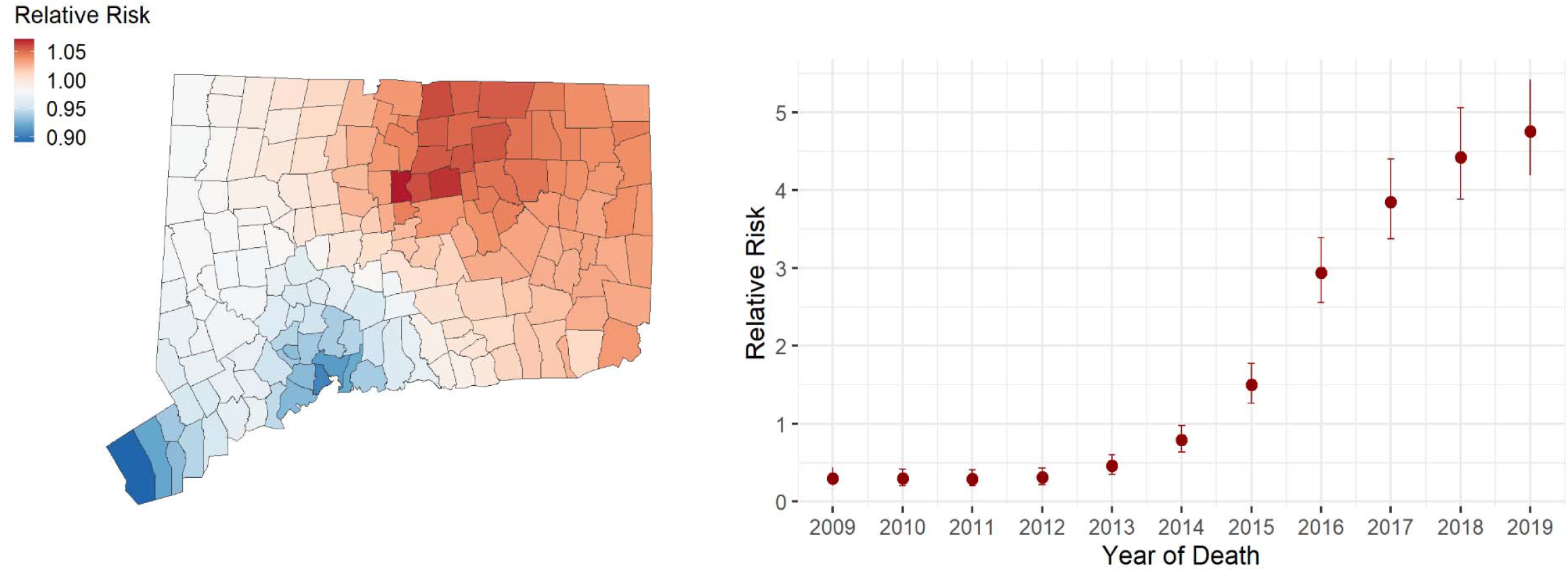
Posterior distributions of estimated parameters for spatial (*α*_*i*_) and temporal (*φ*_*t*_) random effects from the Bayesian space-time Poisson model for fentanyl-detected overdose deaths. The random effects were exponentiated. Larger values of exponentiated coefficients indicate the higher risk of being fentanyl-detected among all opioid overdose death at town level in Connecticut. Note: link to the Connecticut Towns Index Map (https://portal.ct.gov/-/media/DEEP/gis/Resources/IndexTownspdf.pdf)

### Sensitivity Analysis 2

In this sensitivity analysis, we still used the binomial model as described in the main text Methods section. We restricted the analytical sample to adult (aged ≥18) opioid-detected overdose deaths, and modeled the probability of being fentanyl-detected given an adult opioid overdose death in the binomial model. We included the town-level adult population size instead of the town-level total population size as a covariate. As a result, 6618 adult opioid-detected overdose deaths were included in this sensitivity analysis, with 18 (aged <18) deaths excluded (4 were fentanyl-detected). The results were shown in Appendix S6 Figure 2.

**Appendix S6. Figure 2.**
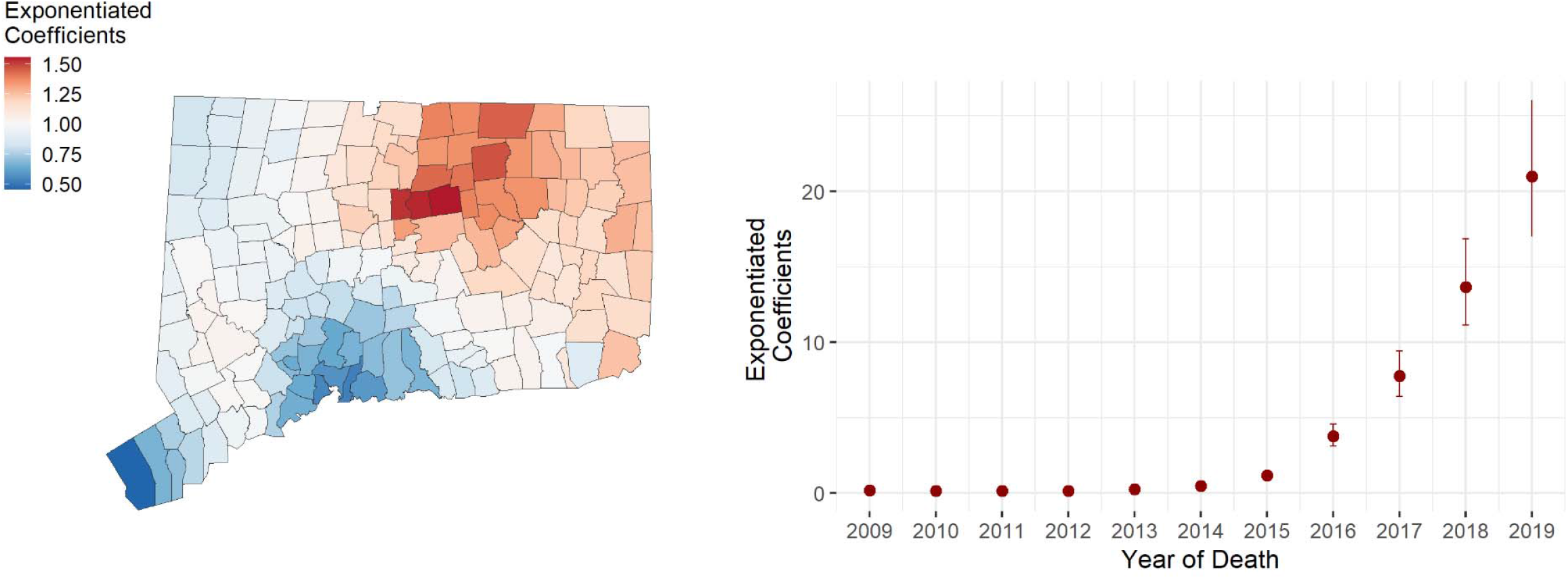
Posterior geometric means of the autoregressive spatial (*µ*_*i*_) and temporal (*φ*_*t*_) random effects from the Bayesian space-time binomial model for adult fentanyl-detected overdose deaths. The left panel depicts the spatial patterns of adult fentanyl-detected overdose deaths when holding temporal terms and other covariates constant. The right panel depicts the temporal patterns of adult fentanyl-detected overdose deaths when holding spatial terms and other covariates constant. The random effects were exponentiated. Larger values of exponentiated coefficients indicate the higher probability being fentanyl detected given an opioid overdose death at town level in Connecticut. Note: link to the Connecticut Towns Index Map (https://portal.ct.gov/-/media/DEEP/gis/Resources/IndexTownspdf.pdf)

### Sensitivity Analysis 3

In this sensitivity analysis, instead of using gamma priors for precision hyperparameters in Bayesian space-time Poisson model for overall opioid-detected overdose deaths, we employed penalized complexity (PC) priors for the prior distribution of hyperparameters. The hyperprior distributions are chosen with the PC framework. We let the precision parameters *τ*_*u*_, *τ*_*v*_, *τ*_*π*_ and *τ*_*ρ*_ (as described in Appendix S1) ∼*PC*(0.2/0.31, 0.01), which corresponds to 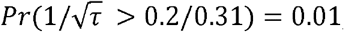, leading to the prior standard deviation being 0.2. The results of this sensitivity analysis were shown in Appendix S6 Figure 3.

**Appendix S6. Figure 3.**
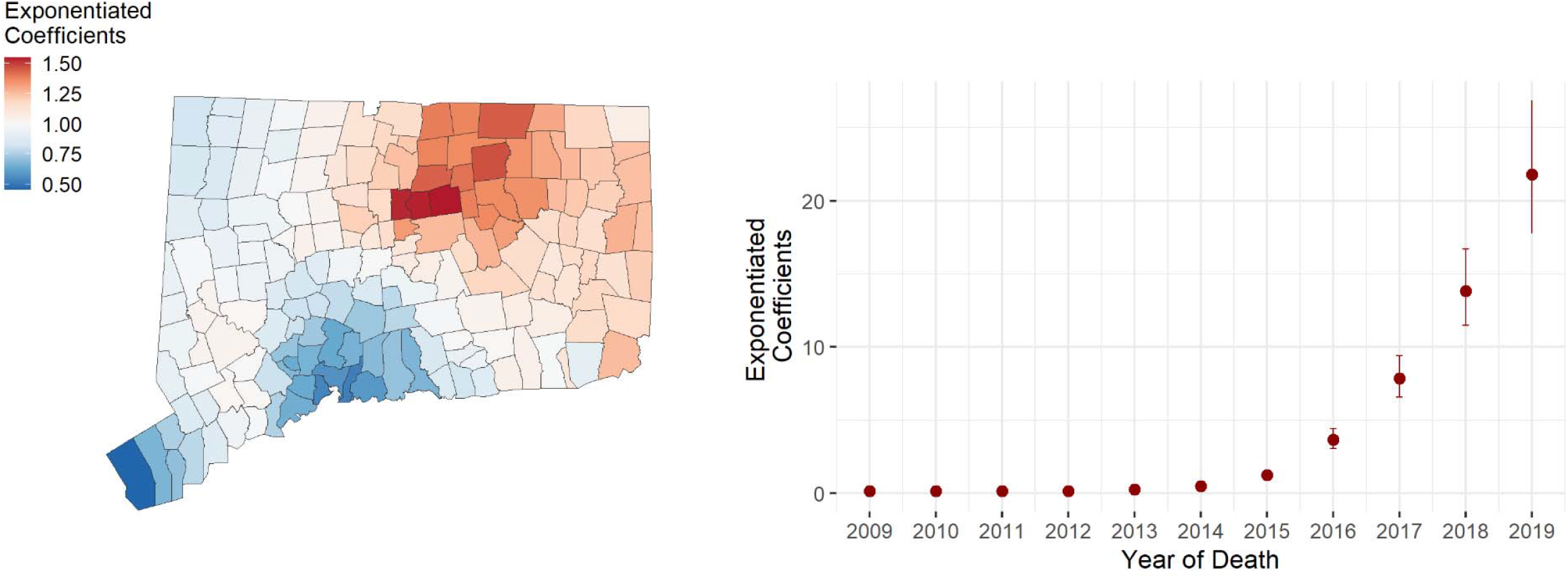
Posterior distributions of estimated parameters for spatial (*µ*_*i*_) and temporal (*φ*_*t*_) random effects from the Bayesian space-time binomial model for fentanyl-detected overdose deaths using penalized complexity priors for hyperparameters. The left panel depicts the spatial patterns of fentanyl-detected overdose deaths when holding temporal terms and other covariates constant. The right panel depicts the temporal patterns of fentanyl-detected overdose deaths when holding spatial terms and other covariates constant. The random effects were exponentiated. Larger values of exponentiated coefficients indicate the higher probability being fentanyl detected given an opioid overdose death at town level in Connecticut. Note: link to the Connecticut Towns Index Map (https://portal.ct.gov/-/media/DEEP/gis/Resources/IndexTownspdf.pdf)

